# Lockdown and non-COVID-19 deaths: Cause-specific mortality during the first wave of the 2020 pandemic in Norway. A population-based register study

**DOI:** 10.1101/2021.02.09.21251326

**Authors:** Guttorm Raknes, Marianne Sørlie Strøm, Gerhard Sulo, Simon Øverland, Mathieu Roelants, Petur Benedikt Juliusson

## Abstract

**Objective:** To explore the potential impact of the first wave of COVID-19 pandemic on all cause and cause-specific mortality in Norway.

**Design:** Population based register study.

**Setting:** The Norwegian cause of Death Registry and the National Population Register of Norway.

**Participants:** All recorded deaths in Norway during March to May from 2010 to 2020.

**Main outcome measures:** Rate (per 100 000) of all-cause mortality and causes of death in the EU Shortlist for Causes of Death March to May 2020. The rates were age-standardised and adjusted to a 100% register coverage and compared with a 95% prediction interval (PI) based on corresponding rates for 2010-2019.

**Results:** 113 710 deaths were included, of which 10 226 from 2020. We did not observe any deviation from predicted total mortality. There were fewer than predicted deaths from chronic lower respiratory diseases excluding asthma (11.4, 95% PI 11.8 to 15.2) and from other non-ischemic, non-rheumatic heart diseases (13.9, 95% PI 14.5 to 20.2). The death rates were higher than predicted for Alzheimer’s disease (7.3, 95% PI 5.5 to 7.3) and diabetes mellitus (4.1, 95% PI 2.1 to 3.4).

**Conclusions:** There was no significant difference in the frequency of the major causes of death in the first wave of the 2020 COVID-19 pandemic in Norway. An increase in diabetes mellitus deaths and reduced mortality due to some heart and lung conditions may be linked to infection control measures.

## INTRODUCTION

The first two Norwegians deaths linked to COVID-19 in Norway were recorded on March 12^th^ 2020. [1] The same day the Norwegian government imposed the most comprehensive infection control measures since World War II (Text box1). [2]

### TEXT BOX1: MEASURES IMPOSED BY THE NORWEGIAN GOVERNMENT ON MARCH 12, 2020 TO LIMIT THE SPREAD OF THE SARS-COV-2 VIRUS

Schools, universities, and kindergartens, shops, and services considered non-essential were shut down

Mandatory 14-day quarantine for all persons entering Norway from countries with a high transmission rate, or after exposure to potential contagion

People were told to isolate themselves and, if possible, to get tested at the slightest sign of respiratory infection

General calls for hand hygiene, to keep a distance of at least two meters in the public space, to work from home whenever possible, and to avoid using public transport

Total mortality figures indicate no excess mortality in Norway in spring 2020. [3] However, total mortality figures do not reflect potential changes in the cause composition of deaths or specific changes for certain causes.

There are concerns that the lockdown and social distancing measures may cause unintended negative consequences for public health and mortality. [4] Some of the specific concerns relate to patients potentially avoiding contact with health services, even for potentially critical symptoms such as chest pain or signs of stroke, [5] or increased suicide rates due to worsening of mental illness in the context of social distancing. [6] On the other hand, social distancing may contribute to fewer infectious diseases in general, such as pneumonia and influenza. The general health of patients with chronic conditions such as heart disease, dementia, and diabetes, may also be more stable in the absence of acute infections, leading to lower hospitalization rates and maybe mortality.

In this study, we used data from the Norwegian Cause of Death Registry to explore whether cause-specific mortality in Norway during the first wave of the COVID-19 pandemic (March to May 2020) was different from that observed during the same months in the preceding years.

## METHODS

We conducted an observational, population-based register study.

Data were obtained from the Norwegian Cause of Death Registry and included information from all death certificates received by December 11^th^, 2020.

The study included all deaths occurring from March 1st until May 31st, 2020, and all deaths from March 1^st^ to May 31^st^ in the years 2010-2019 as reference. The data included Norwegian citizens who died abroad, but not foreign citizens who died in Norway.

Main outcomes were mortality rates (per 100 000 inhabitants) for all items on the 2012 EU Shortlist for Causes of Death. [7] Analyses were performed for all-cause mortality, deaths caused by diseases and external causes, the 17 level-1 causes (including external causes), on all 35 level-2 causes, and all 31 level-3 causes. We present results of analyses for all level 2 and 3 causes of death from the EU shortlist when the observed age-standardised rates were outside the 95% prediction interval. Special attention was given to suicide rates, as it was feared that this rate would increase during the lockdown. For respiratory diseases, we calculated rates both including and excluding COVID-19.

The cause of death is reported by a physician to the Norwegian Cause of Death Registry and the Population Register by completing and mailing an international death certificate (1993 version) [8] or online by submitting an electronic death certificate [9].

The ICD-10 code of the cause of death in the register is determined from the death certificate by an automated coding system (IRIS) that is based on the Automatic Classification of Medical Entry (ACME) software. [10, 11] When needed, the cause of death is manually evaluated by trained staff from the Norwegian Cause of Death Registry. The term “Cause of Death” used throughout this manuscript is equivalent to the “underlying cause of death” as defined by WHO. [12]

### Bias

Although it is mandatory by law to report all deaths to the Norwegian Cause of Death Registry, not all deaths are captured. In some cases, no death certificate is submitted, or some of the involved parties may fail to forward the document to the register. Approximately two percent of death certificates are missing in a typical year. Also, a portion of death certificates is submitted with a delay of several months. Since we retrieved the data several months in advance of the annual publication of the official Norwegian cause of death statistics, bias due to a lower coverage of late incoming certificates is a concern for the 2020 data.

Efforts were made to reduce number of missing causes of death in the 2020 data. The introduction of the digital death certificate contributed to speeding up the logistics, as the register receives electronic certificates within 24 hours. Since the cause of death is more often missing for persons who died abroad, and we also analysed the data excluding these.

We were aware of an increase in the number of deaths at home (outside hospitals and nursing homes), probably secondary to the infection control measures. This could lead to bias, for example due to a larger proportion of death certificates submitted by a general practitioner. Electronic death certificates were available before, during and after the first wave of the pandemic in 2020, and differences between the electronic and paper registration forms could lead to systematic differences in the reporting of cause of death. Before 2020, death certificates were almost exclusively paper-based.

The pandemic itself may also have influenced how doctors determined causes of death on the death certificate. For example, COVID-19 may have changed the level of precision for respiratory causes of death.

### Statistical methods

Age-standardised death rates for the period March-May 2020 were compared with those of the same period during years 2010-2019. Age-standardised mortality rates were computed by the direct standardization method, using 5-year age strata and the European Standard Population of 2013 as standard population. [13] The ‘at risk’ population was defined as the Norwegian population according to Statistics Norway on January 1^st^ of each year. [14]

The age-standardised rates were adjusted to 100% coverage for all years to reduce the impact of missing data when comparing different years. We adjusted each cause of death group, subgroup or diagnose with a proportion of missing data, corresponding to the proportion the outcome made up of the total non-missing data for each year. The death rates used for the analyses are thus age-standardised, adjusted data, but raw rates not adjusted for missing data also represented in figures.

The age-standardised rates were adjusted to 100% coverage for all years to reduce the impact of missing data when comparing different years. We adjusted the rate of each cause of death by multiplying the total number of deaths in a given year with the proportion of deaths due to known causes. The death rates used for the analyses are thus age-standardised, adjusted for missing data, but raw rates not adjusted for missing data also represented in tables and figures.

Observed rates of all cause or disease specific causes of mortality in March – May 2020 were compared with projections based on the same causes during the same months in 2010-2019. Projections were estimated with linear regression and reported as a 95% prediction interval. Rates outside this interval were considered as a statistically significant change. A Durbin-Watson test was used to detect autocorrelation among the annual observations. + summary of findings from this test.

The number of cases with missing cause of death was determined by comparing the number of available death certificates with the total number of deaths according to the National Population Register. We assumed a proportional distribution of causes of death among the missing death certificates. To assess whether missing or delayed death certificates introduced bias, we examined the distribution of age and sex of the deceased and the location of death (home or hospital, nursing home). A similar comparison was made for causes of deaths based on electronic vs. paper-based death certificates.

### Ethics

All analyses were performed on aggregated data, we did not have access to directly or indirectly person identifiable data at any time. Updated data are publicly available from the data repository of the Norwegian Cause of Death Registry. [15]

It is statutory that all deaths that include Norwegian citizens must be registered with the cause of death in the Norwegian Cause of Death Registry. [16] Prior consent to the storage of personal data in the register is not obtained, and it is not possible to opt out. Approval from the ethics committee or privacy ombudsman for research was neither required nor expedient.

## RESULTS

In total, 113 710 deaths were identified in the months March, April, May since 2010. There were 216 cases of COVID-19 as listed cause of death.

Overall, the cause of death was missing in 2 286 cases (2.0%). In 2020 the cause of death was unknown in 568 cases (5.6%), including 131 (out of a total of 133) deaths that occurred abroad.

Patient characteristics for deaths in data are presented in Table 1.

**Table 1.** Characteristics of persons dying during March-May, 2010-2020.

|  | 2010-2019 |  | 2020 |  |
| --- | --- | --- | --- | --- |
| **Mean age (± SD)** | 78.7 | (±15.2) | 78.7 | (±14.8) |
| **Median age (IQR)** | 82 | (17) | 82 | (17) |
| **N (%) females** | 52 910 | (51.1) | 4 909 | (50.8) |
| **N (%) external cause** | 6 394 | (6.3) | 598 | (6.2) |
| **N (%) death at home** | 14 214 | (14.0) | 1 554 | (16.1) |
| **N all deaths** | 103 484 |  | 10 226 |  |

Rates of all causes, diseases, and level-1 EU shortlist causes of death are presented in Table 2 while rates of all (level 1 to 3) EU shortlist causes of death are presented in Supplementary table 1. Observations for all causes, diseases, and external causes are presented in Figure 1, and for the most frequent level 1 causes (cancer, cardiovascular diseases, and respiratory diseases with and without COVID-19) in Figure 2. Causes of death for which the observed age-standardised rate fell outside the prediction interval are presented in Table 3 and in Figure 3. Suicide rates are found in Figure 4. There was a lower than predicted rate for other heart diseases (−19.7%) and other chronic lower respiratory diseases (−13.6%) (Table 3, Figure 2). There was a higher-than-expected rate for diabetes mellitus (49.9%) and Alzheimer’s disease (14.9%) (Table 3, Figure 3). Among the rarer causes of death there was a higher-than-predicted rate for oesophageal cancer, non-malignant neoplasms and certain conditions originating in the perinatal period. The suicide rate did not deviate significantly from the projection based on 2010-2019 (Figure 3). No significant autocorrelation was observed.

**Table 2.** March to May death rates for all chapters in the EU Shortlist for Causes of Death. Death rates are adjusted for missing causes of death, unadjusted rates are also shown. Predicted death rates for 2020 with 95% prediction intervals. All rates are age-standardised. *Respiratory diseases also presented with COVID-19 (officially not in EU shortlist chapter 8).

| EU shortlist chapter | Underlying cause of death | 2010-2019 | 2020 |  |  |  |  |
| --- | --- | --- | --- | --- | --- | --- | --- |
|  |  | Average death rate | N | Predicted death rate | Unadjusted rate | Death rate | 95% prediction interval |
|  | All causes | 238.53 | 1 0226 | 214.86 | 214.38 | 214.38 | (202.42 to 227.30) |
|  | Diseases | 224.48 | 9 280 | 201.12 | 190.48 | 200.96 | (188.93 to 213.31) |
| 1 | Infectious and parasitic diseases | 5.69 | 219 | 5.52 | 4.64 | 4.89 | (4.43 to 6.61) |
| 2 | Neoplasms | 65.25 | 2 695 | 58.46 | 56.13 | 59.25 | (55.94 to 60.98) |
| 3 | Diseases of the blood and blood-forming organs and certain disorders involving the immune mechanism | 0.83 | 36 | 0.61 | 0.76 | 0.80 | (0.31 to 0.92) |
| 4 | Endocrine, nutritional, and metabolic diseases | 5.75 | 295 | 5.22 | 6.18 | 6.52 | (4.24 to 6.20) |
| 5 | Mental and behavioural disorders | 14.76 | 775 | 18.29 | 16.55 | 17.45 | (15.5 to 21.07) |
| 6 | Diseases of the nervous system and the sense organs | 11.23 | 625 | 13.07 | 13.22 | 13.94 | (11.78 to 14.35) |
| 7 | Diseases of the circulatory system | 70.31 | 2 182 | 50.53 | 46.03 | 48.55 | (46.18 to 54.89) |
| 8 | Diseases of the respiratory system (excl. COVID-19) | 26.49 | 1 005 | 27.53 | 21.29 | 22.45 | (22.95 to 32.11) |
| * | Diseases of the respiratory system (incl. COVID-19) | 26.49 | 1 221 | 27.53 | 25.93 | 27.34 | (22.95 to 32.11) |
| * | COVID-19 | 0.00 | 216 | 0.00 | 4.64 | 4.89 | (0.00 to 0.00) |
| 9 | Diseases of the digestive system | 7.45 | 302 | 6.98 | 6.34 | 6.69 | (5.92 to 8.03) |
| 10 | Diseases of the skin and subcutaneous tissue | 0.60 | 27 | 0.79 | 0.58 | 0.61 | (0.48 to 1.10) |
| 11 | Diseases of the musculoskeletal system/connective tissue | 1.62 | 76 | 1.97 | 1.60 | 1.69 | (1.49 to 2.45) |
| 12 | Diseases of the genitourinary system | 4.99 | 191 | 4.37 | 4.04 | 4.26 | (3.04 to 5.71) |
| 13 | Complications of pregnancy, childbirth and puerperium | 0.01 | 0 | 0.00 | 0.00 | 0.00 | (-0.02 to 0.03) |
| 14 | Certain conditions originating in the perinatal period | 0.35 | 22 | 0.18 | 0.40 | 0.43 | (0.00 to 0.36) |
| 15 | Congenital malformations and chromosomal abnormalities | 0.63 | 37 | 0.65 | 0.71 | 0.75 | (0.44 to 0.85) |
| 16 | Symptoms, signs, ill-defined causes | 8.53 | 357 | 6.94 | 7.38 | 7.79 | (4.06 to 9.82) |
| 17 | External causes of morbidity and mortality | 13.99 | 598 | 13.63 | 12.07 | 12.76 | (11.08 to 16.18) |

**Table 3:** Observed and predicted mortality from. causes of death where the death rate of March to May 2020 deviated statistically significantly from the 95% prediction interval. Sorted descending after number of deaths. Death rates are adjusted for missing causes of death, unadjusted rates are also shown. All rates are age-standardised.

| EU Shortlist chapter | Underlying cause of death | 2010-19 | 2020 |  |  |  |  |
| --- | --- | --- | --- | --- | --- | --- | --- |
|  |  | Average death rate | N | Predicted death rate | Unadjusted death rate | Death rate | 95% prediction interval |
| 7.2 | Other heart diseases | 20.16 | 622 | 17.33 | 13.20 | 13.92 | (14.50 to 20.17) |
| 8.3.2 | *Chronic lower respiratory diseases (excl. Asthma)* | 13.08 | 509 | 13.48 | 10.79 | 11.38 | (11.80 to 15.15) |
| 6.2 | Alzheimer's disease | 5.23 | 323 | 6.36 | 6.94 | 7.31 | (5.47 to 7.26) |
| 4.1 | Diabetes mellitus | 3.64 | 185 | 2.76 | 3.92 | 4.13 | (2.10 to 3.42) |
| 2.2 | Non-malignant neoplasms (benign and uncertain) | 1.52 | 87 | 1.37 | 1.84 | 1.94 | (0.80 to 1.93) |
| 2.1.2 | *Malignant neoplasm of oesophagus* | 1.22 | 74 | 1.14 | 1.51 | 1.60 | (0.72 to 1.56) |
| 14 | **Certain conditions originating in the perinatal period** | **0.35** | **22** | **0.18** | **0.40** | **0.43** | **(0.00 to 0.36)** |

**Figure 1:**
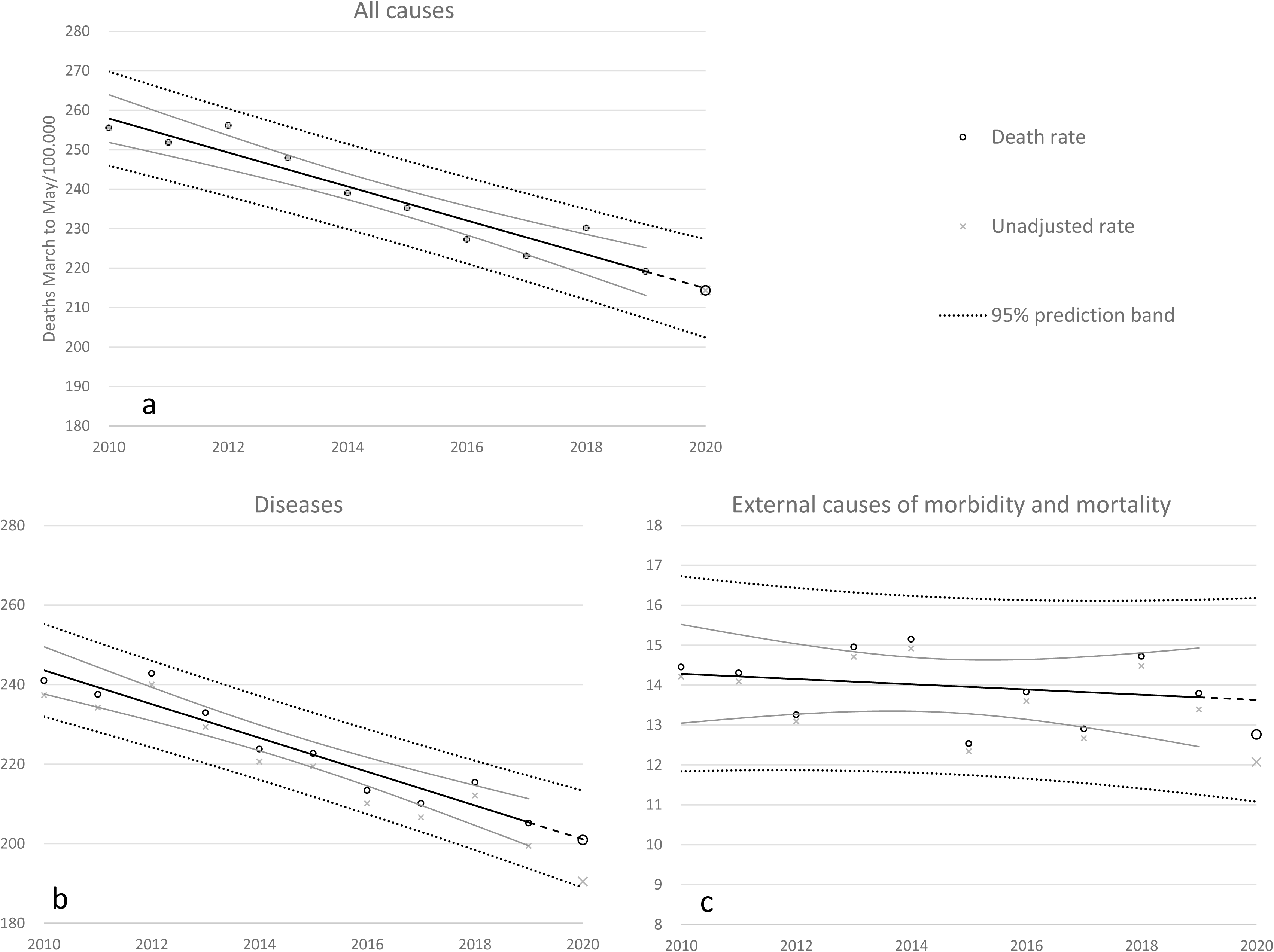
Observed and predicted death rates (linear) with 95% confidence bands for all-cause mortality (panel a), total mortality excluding external causes (panel b) and mortality due to external causes (panel c).

**Figure 2:**
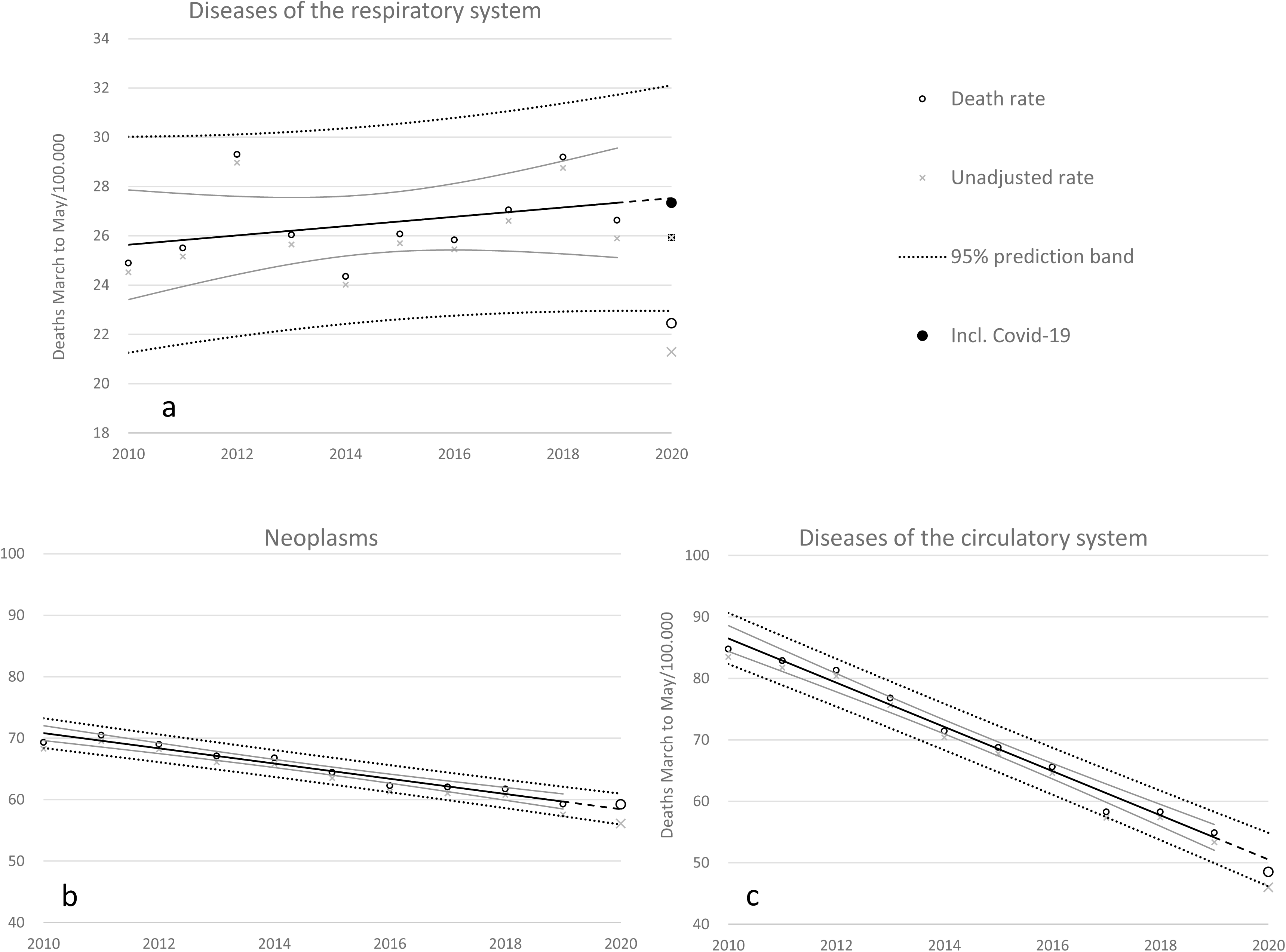
Observed and predicted death rates (linear) with 95% confidence bands for respiratory diseases (panel a), neoplasms (panel b), and cardiovascular diseases causes (panel c). For 2020, mortality from respiratory diseases is presented both with and without COVID-19.

**Figure 3:**
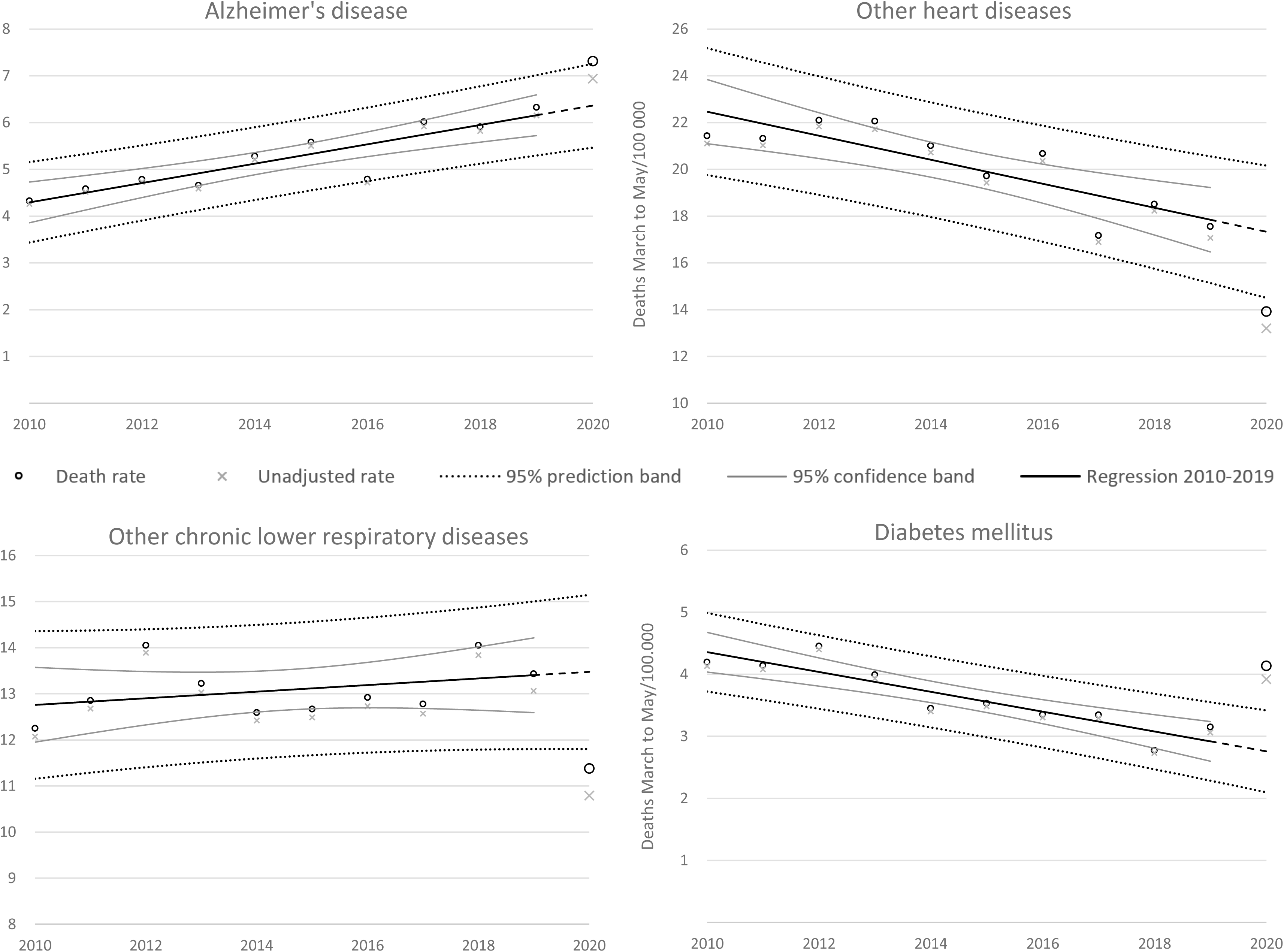
Observed and predicted death rates (linear) with 95% confidence bands for causes of death with higher of lower rates than expected.

**Figure 4:**
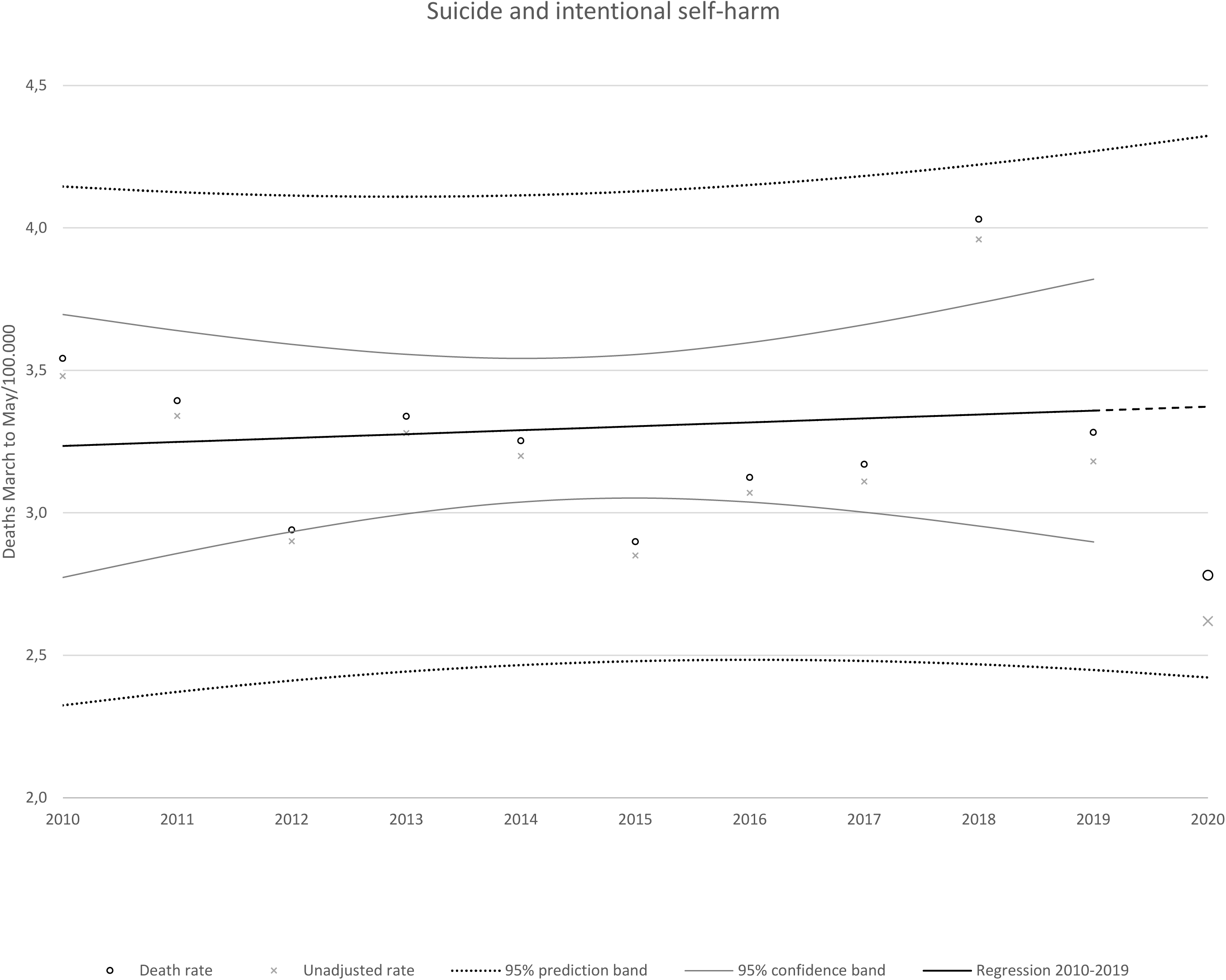
Observed and predicted death rate (linear) with 95% confidence bands for suicide and intentional self-harm.

Characteristics of persons with or without known cause of death, death at home and death elsewhere, and cause of deaths reported electronically or with a paper-based death certificate are presented in Supplementary table 2. Age and sex distribution were almost identical in cases with or without known cause of death, and in digital and paper-based death certificates. Among deaths at home, median age was nine years lower, there was a lower proportion females and external causes of death were more frequent.

## DISCUSSION

### Principal findings

In this analysis of mortality during the first wave of the COVID-19 pandemic in Norway we observed no excess mortality and no substantial changes in the rates for neoplasms, cardiovascular disease, suicide, or other external causes. There was a lower than predicted death rate for respiratory diseases, which is probably attributable to lower mortality from chronic lung diseases (excluding asthma). The mortality rate was also lower than predicted for other non-ischemic heart diseases (excluding valvular and rheumatic heart conditions) and for Alzheimer’s disease. The observed death rate of diabetes mellitus was 45% higher than predicted. Within neoplasms, we observed higher than expected rates for oesophageal cancer and benign neoplasms.

### Strengths and imitations

The Cause of Death Registry represent the main source of information for monitoring mortality. The Registry is based on established international coding practices that offer comparability and can be used to examine trends over time. The Norwegian Cause of Death Registry covers all deaths in Norway, and since it is possible to verify and validate its content using other sources, it is regarded as highly reliable. [17] As in the other Nordic countries, the Norwegian society is characterized by a high degree of trust between citizens and in the authorities. As a result, there is evidence of a high degree of compliance with governmental measures during the pandemic, which has probably contributed to low coronavirus transmission and death rates during the pandemic. [18] There is thus reason to believe that Norway is well suited for studies on the effects of population-based infection control measures.

Although death registers typically cover the whole population, they are prone to bias due to incomplete information on individual cases. Our main concern was bias due to loss or late incoming death certificates. However, as shown in Supplementary table 2, descriptive characteristics indicate that the group of patients with or without known cause of death was not systematically different.

The proportion of deaths at home from March to May 2020 was 15% higher than in the same months in 2010-2019. Both age and the proportion of females were considerably lower among deaths at home in 2020 (Supplementary table 2), but this was also true in 2010-19 (data not shown). The absolute increase in deaths at home (200-250 estimated deaths) should not be sufficient to skew the findings significantly. Differences in patient characteristics from electronic and paper-based death certificates were also marginal, and thus unlikely to be a significant source of bias.

Doctors often have limited information about the patient when recording the cause of death on a death certificate, and a subjective assessment may occur. There may have been some systematic changes in the diagnostic practice and determination of the cause of death during the pandemic, especially when it comes to respiratory diseases. A more frequent use of chest x-rays may have biased the rates of the different respiratory causes of death. These effects are difficult to quantify, but we do not believe that the resulting bias is clinically relevant for most causes of death.

Another limitation is the large and diverse number of outcomes. We examined 83 EU shortlist causes of deaths without correcting for multiple testing. Even under normal circumstances it is therefore likely that some rates would fall outside the expected prediction interval. We could have used wider prediction intervals to adjust for multiple comparisons and avoid false positive findings, but this would at the same time increase the probability of a false negative finding. A comparison of the causes of death in Table 3 with a 99% prediction interval, shows that only diabetes mellitus remains significantly outside the predicted range. This means that great care should be taken when interpreting the results.

### Strengths & weaknesses in relation to other studies & key differences

We have been unable to identify other studies with similar focus. The SARS-COV-2 virus was only identified in December 2019, and access to high-quality data with sufficient detail in such a short time is limited. Some studies reported on all-cause mortality, [3,19] in relation to COVID-19 and infection control measures, but data on specific causes of death is usually lacking.

### Possible mechanisms & explanations for findings

Only 216 Norwegians died from COVID-19 in March to May 2020, and the infection rate was also very low. Reports from the Norwegian National Institute of Public Health even showed a lower-than-expected mortality in some of the weeks in the first wave of the pandemic, [20] indicating that misclassification of COVID-19 deaths was almost non-existent. It is therefore unlikely that SARS-COV-2 infections had a significant impact on the mortality of other causes in Norway in this period.

The unexpected low death rate from chronic lung diseases could be a consequence of the anti-pandemic measures. For instance, exacerbations from chronic obstructive pulmonary disease (COPD) are often caused by respiratory infections. Social distancing and improved hygiene may have been protective, even for non-respiratory diseases. The acute myocardial infarction death rate was the lowest since recordings started in 1970, but this fits into a decades long downward trend. In contrast, the number of deaths due to Alzheimer’s disease has increased since the early 1990s. The observed mortality rate for Alzheimer’s disease in 2020 was slightly higher than the upper limit of the prediction interval. Many Alzheimer’s patients live in nursing homes where infection control measures were extra strict. There have been concerns that this patient group may have problems complying with the infection control measures, and that basic needs of vulnerable patients have been set aside. [21]

The most unexpected and striking finding was the large and significant increase in diabetes mellitus as cause of death (4.1 deaths per 100 000). The increase was 45% compared to the predicted rate, which is equivalent to an excess of 74 deaths in March to May 2020. One explanation could be that some diabetes patients inappropriately avoided contact with healthcare professionals, which may have contributed to poor blood glucose control. A recent survey among diabetes nurses across Europe confirmed that there are significant physical and psychological problems in their patient populations during COVID-19, and that clinical diabetes services have been disrupted. [22] We were not able to differentiate between type 1 and type 2 diabetes.

The observed increase in deaths from oesophageal cancer, benign neoplasms and perinatal conditions should be followed closely in the time ahead. However, we find it unlikely that these findings are associated with infection control measures because there is no obvious underlying mechanism. These causes are also rare, and therefore more prone to become outliers in in a study with many outcomes. The perinatal causes were 140% higher than predicted, but there were only 22 deaths. Deaths due to oesophageal cancer end non-malignant neoplasms were only marginally higher than the upper limit of the 95% prediction interval.

We did not find an increased suicide rate, but the observation window of our study is limited to the first three months of the pandemic in Norway. It is possible that negative effects of the pandemic and social distancing measures will have more impact over time. So far, COVID-19 is coded separately in the cause of death register, and not as an EU shortlist chapter 8 respiratory disease. As seen in Figure 2, respiratory disease as cause of death is within the 95% prediction interval when including COVID-19.

### Potential implications for clinicians or policymakers

This study did not reveal an alarming increase in the causes of death that could suggest unacceptable negative effects of the infection control measures on public health or mortality, including suicide.

The clinical relevance of the observed changes in cause of death rate is uncertain. Diabetes mellitus may be a notable exception, and extra efforts should be made to monitor diabetes mortality and maybe even improve diabetes care throughout the pandemic.

### Unanswered questions and future research

The current pandemic is caused by a virus that was only discovered in December 2019. We have currently only examined mortality during the first wave in Norway, but similar continuous monitoring is warranted until the population is sufficiently vaccinated. Results from this study highlight the importance to closely monitor morbidity and mortality due to diabetes mellitus in particular.

## Data Availability

All analyses were performed on aggregated data, we did not have access to directly or indirectly person identifiable data at any time.
The main analyses are based on cause of death data that are available from the Norwegian Cause of Death Registry (http://statistikkbank.fhi.no/dar/), a public, open access repository (in Norwegian only). The repository is updated regularly, and late incoming death certificates provide a gradually improving coverage. The analyses in this study are based on data retrieved on December 11, 2020; these data are found in in Supplementary table 3. 
Data on the number of death certificates submitted electronically are not publicly available, but these will be made available on request. Weekly death statistics from the National Population Register can be accessed via Statistics Norway's webpages (https://www.ssb.no/befolkning/artikler-og-publikasjoner/her-finner-du-ukentlige-tall-pa-antall-dode). 

http://statistikkbank.fhi.no/dar/

https://www.ssb.no/befolkning/artikler-og-publikasjoner/her-finner-du-ukentlige-tall-pa-antall-dode

## DECLARATIONS

## Acknowledgements

We would like to thank Olaug Margrethe Askeland and Gunhild Forland Slungård for preparing the data files at short notice.

## Copyright/licensing

The Corresponding Author has the right to grant on behalf of all authors and does grant on behalf of all authors, an exclusive non exclusive licence on a worldwide basis to the BMJ Publishing Group Ltd to permit this article (if accepted) to be published in BMJ editions and any other BMJPGL products and sublicences such use and exploit all subsidiary rights, as set out in our licence.

## Author contributions

All authors contributed substantially to the conception and design of the study, to the interpretation of data, to critical revision of the manuscript’s intellectual content, and approved the final version. GR (the guarantor) performed the analyses and drafted the manuscript.

The authors agree to be accountable for all aspects of the work in ensuring that questions related to the accuracy or integrity of any part of the work are appropriately investigated and resolved.

## Transparency declaration

The lead author (guarantor of manuscript) affirms that this manuscript is an honest, accurate, and transparent account of the study being reported; that no important aspects of the study have been omitted; and that any discrepancies from the study as planned (and, if relevant, registered) have been explained.

## Patient and public involvement statement

The study was partly initiated due to questions from the press and public. Patients and the public were not involved in the development and conduct of the study or interpretation or presentation of the results. It is not possible for either researchers or the general population to influence which parameters are registered in the Norwegian Cause of Death Registry. All data are freely available on request.

## Data sharing statement

All analyses were performed on aggregated data, we did not have access to directly or indirectly person identifiable data at any time.

The main analyses are based on cause of death data that are available from the Norwegian Cause of Death Registry (http://statistikkbank.fhi.no/dar/), a public, open access repository (in Norwegian only). The repository is updated regularly, and late incoming death certificates provide a gradually improving coverage. The analyses in this study are based on data retrieved on December 11, 2020; these data are found in in Supplementary table 3.

Data on the number of death certificates submitted electronically are not publicly available, but these will be made available on request. Weekly death statistics from the National Population Register can be accessed via Statistics Norway’s webpages (https://www.ssb.no/befolkning/artikler-og-publikasjoner/her-finner-du-ukentlige-tall-pa-antall-dode).

## Reporting guideline

A filled in STROBE checklist for cross sectional studies is submitted. We are aware that this is not intended for population based register studies, but most items in the STROBE checklist are relevant.

## Protocol

Not applicable

## Patient consent

Not applicable.

The article has not been submitted to any other journal previously.

## SUPPLEMENTARY MATERIAL

**Supplementary table 1:** Observed and predicted mortality rates with 95% prediction intervals for all analysed items in the EU Shortlist for Causes of Death and deaths from COVID-19. Diseases and level 1 (EU shortlist main chapters) in bold. Level 3 in italics. Rates (per 100 000) are age-standardised. Predictions are based on age-standardised rates adjusted to 100% coverage, adjusting for cases with unknown cause of death. *Respiratory diseases also presented with COVID-19 (officially not in EU shortlist chapter 8).

**Supplementary table 2**: Characteristics of persons (2020) with or without known cause of death, death at home and death elsewhere, and cause of deaths reported digitally or with a paper-based death certificate. Supplementary file 3: Underlying data. Cause specific deaths March to May 2010 to 2020 with rates recorded in the Norwegian Cause of Death Registry by 11 December 2020. The causes have been grouped according to the EU Shortlist for Causes of Disease. Level 1 corresponds to the 17 main chapter

**Supplementary table 3:** Underlying data. Cause specific deaths March to May 2010 to 2020 (column E to O) with rates (per 100000, column P to Z) recorded in the Norwegian Cause of Death Registry by 11 December 2020. The causes have been grouped according to the EU Shortlist for Causes of Disease. Level 1 corresponds to the 17 main chapters. *Respiratory diseases also presented with COVID-19 (officially not in EU shortlist chapter 8).

