## Supplementary table 1 for "Lockdown and non-COVID-19 deaths: Cause-specific mortality during the first wave of the 2020 pandemic in Norway. A population-based register study"

**Supplementary table 1:** Observed and predicted mortality rates with 95% prediction intervals for all analysed items in the EU Shortlist for Causes of Death and deaths from COVID-19. Diseases and level 1 (EU shortlist main chapters) in bold. Level 3 in italics. Rates (per 100 000) are age-standardised. Predictions are based on age-standardised rates adjusted to 100% coverage, adjusting for cases with unknown cause of death. *Respiratory diseases also presented with COVID-19 (officially not in EU shortlist chapter 8).

| | | **2010-2019** | **2020** | | | | |
|---|---|---|---|---|---|---|---|
| **EU shortlist chapter** | **Underlying cause of death** | **Average death rate 2010-19** | **N** | **Predicted death rate** | **Unadjusted rate** | **Death rate** | **95% prediction interval** |
| | All causes | 238.53 | 1 0226 | 214.86 | 214.38 | **214.38** | (202.42 to 227.30) |
| | Diseases | 224.48 | 9280 | 201.12 | 190.48 | **200.96** | (188.93 to 213.31) |
| **1** | **Infectious and parasitic diseases** | 5.69 | 219 | 5.52 | 4.64 | **4.89** | (4.43 to 6.61) |
| 1.1 | Tuberculosis | 0.12 | 3 | 0.08 | 0.06 | **0.06** | (-0.03 to 0.18) |
| 1.2 | AIDS (HIV-disease) | 0.06 | 4 | 0.05 | 0.08 | **0.08** | (-0.03 to 0.13) |
| 1.3 | Viral hepatitis | 0.07 | 6 | 0.08 | 0.12 | **0.13** | (-0.01 to 0.17) |
| 1.4 | Other infectious and parasitic diseases | 5.44 | 206 | 5.31 | 4.38 | **4.62** | (4.16 to 6.45) |
| **2** | **Neoplasms** | 65.25 | 2695 | 58.46 | 56.13 | **59.25** | (55.94 to 60.98) |
| **2.1** | **Malignant neoplasms** | 63.73 | 2608 | 57.09 | 54.29 | **57.31** | (54.29 to 59.89) |
| *2.1.1* | *Malignant neoplasm of lip, oral cavity, pharynx* | 0.84 | 44 | 0.88 | 0.92 | **0.97** | (0.48 to 1.29) |
| *2.1.2* | *Malignant neoplasm of oesophagus* | 1.22 | 74 | 1.14 | 1.51 | **1.60** | (0.72 to 1.56) |
| *2.1.3* | *Malignant neoplasm of stomach* | 1.85 | 69 | 1.43 | 1.44 | **1.52** | (0.97 to 1.89) |
| *2.1.4* | *Malignant neoplasm of colon, rectum and anus* | 9.42 | 343 | 8.23 | 7.15 | **7.55** | (7.13 to 9.33) |
| *2.1.5* | *Malignant neoplasm of liver and intrahepatic bile ducts* | 1.52 | 82 | 1.72 | 1.69 | **1.78** | (1.07 to 2.36) |
| *2.1.6* | *Malignant neoplasm of pancreas* | 4.09 | 187 | 4.22 | 3.89 | **4.11** | (3.53 to 4.92) |
| *2.1.7* | *Malignant neoplasm of larynx* | 0.22 | 7 | 0.12 | 0.14 | **0.15** | (-0.06 to 0.29) |
| *2.1.8* | *Malignant neoplasm of trachea, bronchus, lung* | 13.26 | 543 | 11.50 | 11.27 | **11.90** | (10.34 to 12.66) |
| *2.1.9* | *Malignant melanoma of skin* | 1.83 | 73 | 1.54 | 1.52 | **1.60** | (1.05 to 2.03) |
| *2.1.10* | *Malignant neoplasm of breast* | 3.61 | 142 | 3.25 | 2.93 | **3.09** | (2.88 to 3.61) |
| *2.1.11* | *Malignant neoplasm of cervix uteri* | 0.42 | 20 | 0.51 | 0.40 | **0.42** | (0.25 to 0.77) |
| *2.1.12* | *Malignant neoplasm of other and unspecified parts of uterus* | 0.87 | 24 | 0.73 | 0.50 | **0.53** | (0.24 to 1.21) |
| *2.1.13* | *Malignant neoplasm of ovary* | 1.68 | 57 | 1.55 | 1.16 | **1.23** | (1.10 to 1.99) |
| *2.1.14* | *Malignant neoplasm of prostate* | 5.89 | 222 | 4.76 | 4.75 | **5.01** | (3.03 to 6.49) |
| *2.1.15* | *Malignant neoplasm of kidney* | 1.55 | 62 | 1.26 | 1.28 | **1.35** | (0.70 to 1.83) |
| *2.1.16* | *Malignant neoplasm of bladder* | 1.82 | 74 | 1.72 | 1.55 | **1.64** | (1.25 to 2.19) |
| *2.1.17* | *Malignant neoplasm of brain and central nervous system* | 1.79 | 80 | 1.69 | 1.59 | **1.68** | (1.20 to 2.19) |
| *2.1.18* | *Malignant neoplasm of thyroid* | 0.18 | 8 | 0.21 | 0.17 | **0.18** | (-0.17 to 0.58) |
| *2.1.19* | *Hodgkin disease and lymphomas* | 1.89 | 89 | 1.54 | 1.87 | **1.97** | (0.68 to 2.39) |
| *2.1.20* | *Leukaemia* | 2.00 | 73 | 2.05 | 1.54 | **1.62** | (0.93 to 3.17) |
| *2.1.21* | *Other malignant neoplasm of lymphoid and haematopoietic tissue* | 1.63 | 57 | 1.45 | 1.19 | **1.26** | (1.07 to 1.82) |
| *2.1.22* | *Other malignant neoplasms* | 6.14 | 278 | 5.61 | 5.83 | **6.15** | (4.54 to 6.68) |
| 2.2 | **Non-malignant neoplasms (benign and uncertain)** | 1.52 | 87 | 1.37 | 1.84 | **1.94** | (0.80 to 1.93) |
| **3** | **Diseases of the blood and blood-forming organs and certain disorders involving the immune mechanism** | 0.83 | 36 | 0.61 | 0.76 | **0.80** | (0.31 to 0.92) |
| **4** | **Endocrine, nutritional and metabolic diseases** | 5.75 | 295 | 5.22 | 6.18 | **6.52** | (4.24 to 6.20) |
| 4.1 | Diabetes mellitus | 3.64 | 185 | 2.76 | 3.92 | **4.13** | (2.10 to 3.42) |
| 4.2 | Other endocrine, nutritional and metabolic diseases | 2.11 | 110 | 2.46 | 2.26 | **2.39** | (1.92 to 3.00) |
| **5** | **Mental and behavioural disorders** | **14.76** | **775** | **18.29** | **16.55** | **17.45** | **(15.50 to 21.07)** |
| 5.1 | Dementia | 13.15 | 697,0 | 16.96 | 14.94 | **15.75** | (14.79 to 19.13) |
| 5.2 | Alcohol abuse (including alcoholic psychosis) | 0.83 | 35,0 | 0.53 | 0.71 | **0.75** | (-0.05 to 1.11) |
| 5.3 | Drug dependence, toxicomania | 0.10 | 7,0 | 0.06 | 0.13 | **0.14** | (-0.07 to 0.19) |
| 5.4 | Other mental and behavioural disorders | 0.68 | 36 | 0.73 | 0.77 | **0.81** | (0.32 to 1.15) |
| **6** | **Diseases of the nervous system and the sense organs** | 11.23 | 625 | 13.07 | 13.22 | **13.94** | (11.78 to 14.35) |
| 6.1 | Parkinson's disease | 2.21 | 119 | 3.11 | 2.54 | **2.68** | (2.30 to 3.91) |
| 6.2 | Alzheimer's disease | 5.23 | 323 | 6.36 | 6.94 | **7.31** | (5.47 to 7.26) |
| 6.3 | Other diseases of the nervous system and the sense organs | 3.79 | 183 | 3.60 | 3.74 | **3.95** | (2.56 to 4.64) |
| **7** | **Diseases of the circulatory system** | 70.31 | 2182 | 50.53 | 46.03 | **48.55** | (46.18 to 54.89) |
| 7.1 | Ischaemic heart diseases | 25.44 | 713 | 15.16 | 14.99 | **15.81** | (13.25 to 17.07) |

| | | | | | | | |
|---|---|---|---|---|---|---|---|
| *7.1.1* | *Acute myocardial infarction* | 15.66 | 380 | 8.35 | 8.02 | **8.46** | (6.97 to 9.74) |
| *7.1.2* | *Other ischaemic heart diseases* | 9.78 | 333 | 6.81 | 6.97 | **7.36** | (5.42 to 8.19) |
| 7.2 | Other heart diseases | 20.16 | 622 | 17.33 | 13.20 | **13.92** | (14.50 to 20.17) |
| 7.3 | Cerebrovascular diseases | 16.22 | 529 | 10.59 | 11.17 | **11.78** | (8.95 to 12.22) |
| 7.4 | Other diseases of the circulatory system | 8.49 | 318 | 7.45 | 6.67 | **7.04** | (6.32 to 8.58) |
| **8** | **Diseases of the respiratory system (excl covid)** | 26.49 | 1005 | 27.53 | 21.29 | **22.45** | (22.95 to 32.11) |
| * | Diseases of the respiratory system (incl covid) | 26.49 | 1221 | 27.53 | 25.93 | **27.34** | (22.95 to 32.11) |
| 8.1 | Influenza | 1.02 | 35 | 2.19 | 0.72 | **0.76** | (0.60 to 3.77) |
| 8.2 | Pneumonia | 9.25 | 302 | 8.58 | 6.40 | **6.75** | (6.20 to 10.97) |
| **8.3** | Chronic lower respiratory diseases | 13.60 | 541 | 13.99 | 11.47 | **12.10** | (12.29 to 15.69) |
| *8.3.1* | *Asthma* | 0.52 | 32 | 0.51 | 0.68 | **0.72** | (0.26 to 0.77) |
| *8.3.2* | *Other chronic lower respiratory diseases* | 13.08 | 509 | 13.48 | 10.79 | **11.38** | (11.8 to 15.15) |
| 8.4 | Other diseases of the respiratory system | 2.62 | 127 | 2.77 | 2.70 | **2.85** | (2.06 to 3.48) |
| **9** | **Diseases of the digestive system** | 7.45 | 302 | 6.98 | 6.34 | **6.69** | (5.92 to 8.03) |
| 9.1 | Ulcer of stomach, duodenum and jejunum | 0.95 | 21 | 0.67 | 0.43 | **0.45** | (0.44 to 0.91) |
| 9.2 | Cirrhosis, fibrosis and chronic hepatitis | 1.14 | 50 | 1.13 | 1.02 | **1.08** | (0.63 to 1.62) |
| 9.3 | Other diseases of the digestive system | 5.36 | 231 | 5.18 | 4.89 | **5.16** | (3.91 to 6.45) |
| **10** | **Diseases of the skin and subcutaneous tissue** | 0.60 | 27 | 0.79 | 0.58 | **0.61** | (0.48 to 1.10) |
| **11** | **Diseases of the musculoskeletal system/connective tissue** | 1.62 | 76 | 1.97 | 1.60 | **1.69** | (1.49 to 2.45) |
| 11.1 | Rheumatoid arthritis and osteoarthrosis | 0.42 | 15 | 0.60 | 0.32 | **0.34** | (0.28 to 0.93) |
| 11.2 | Other diseases of the musculoskeletal system/connective tissue | 1.20 | 61 | 1.37 | 1.28 | **1.35** | (1.11 to 1.62) |
| **12** | **Diseases of the genitourinary system** | 4.99 | 191 | 4.37 | 4.04 | **4.26** | (3.04 to 5.71) |
| 12.1 | Diseases of kidney and ureter | 3.08 | 131 | 2.68 | 2.77 | **2.92** | (1.55 to 3.81) |
| 12.2 | Other diseases of the genitourinary system | 1.91 | 60 | 1.70 | 1.27 | **1.34** | (1.26 to 2.13) |
| **13** | **Complications of pregnancy, childbirth and puerperium** | 0.01 | 0 | 0.00 | 0.00 | **0.00** | (-0.02 to 0.03) |
| **14** | **Certain conditions originating in the perinatal period** | 0.35 | 22 | 0.18 | 0.40 | **0.43** | (0.00 to 0.36) |
| **15** | **Congenital malformations and chromosomal abnormalities** | 0.63 | 37 | 0.65 | 0.71 | **0.75** | (0.44 to 0.85) |
| **16** | **Symptoms, signs, ill-defined causes** | 8.53 | 357 | 6.94 | 7.38 | **7.79** | (4.06 to 9.82) |
| 16.1 | Sudden infant death syndrome | 0.04 | 0 | 0.03 | 0.00 | **0.00** | (-0.06 to 0.12) |
| 16.2 | Unknown and unspecified causes | 5.53 | 254 | 4.63 | 5.24 | **5.53** | (1.94 to 7.33) |
| 16.3 | Other symptoms, signs, ill-defined causes | 2.96 | 103 | 2.28 | 2.14 | **2.26** | (1.17 to 3.38) |
| **17** | **External causes of morbidity and mortality** | 13.99 | 598 | 13.63 | 12.07 | **12.76** | (11.08 to 16.18) |
| 17.1 | Accidents | 10.54 | 452 | 10.13 | 9.32 | **9.84** | (8.27 to 12.00) |
| *17.1.1* | *Transport accidents* | 0.89 | 35 | 0.62 | 0.67 | **0.71** | (0.36 to 0.89) |
| *17.1.2* | *Accidental falls* | 3.41 | 161 | 4.55 | 3.46 | **3.65** | (2.62 to 6.47) |
| *17.1.3* | *Drowning and accidental submersion* | 0.27 | 13 | 0.24 | 0.25 | **0.27** | (-0.05 to 0.53) |
| *17.1.4* | *Accidental poisoning* | 1.69 | 79 | 1.83 | 1.50 | **1.59** | (1.39 to 2.28) |
| *17.1.5* | *Other accidents* | 4.28 | 164 | 2.89 | 3.44 | **3.63** | (-0.09 to 5.87) |
| 17.2 | Suicide and intentional self-harm | 3.30 | 139 | 3.37 | 2.62 | **2.78** | (2.42 to 4.32) |
| 17.3 | Homicide, assault | 0.12 | 6 | 0.07 | 0.11 | **0.12** | (-0.13 to 0.28) |
| 17.4 | Events of undetermined intent | 0.03 | 0 | 0.03 | 0.00 | **0.00** | (-0.06 to 0.13) |
| 17.5 | Other external causes of injury and poisoning | 0.01 | 1 | 0.01 | 0.02 | **0.02** | (-0.04 to 0.07) |
| | **COVID-19** | 0.00 | 216 | 0.00 | 4.64 | **4.89** | (0.00 to 0.00) |
