## Supplementary table 2 for "Lockdown and non-COVID-19 deaths: Cause-specific mortality during the first wave of the 2020 pandemic in Norway. A population-based register study"

**Supplementary Table 2:** Characteristics of persons (2020) with or without known cause of death, death at home and death elsewhere, and cause of deaths reported digitally or with a paper-based death certificate.

| | Cause of death | | | | | Place of death | | | | | Death certificate type | | | |
|---|---|---|---|---|---|---|---|---|---|---|---|---|---|---|
| | Known | | Unknown | | | At home | | All other | | | Digital | | Paper | |
| **N (%)*** | 9 658 | (94.4) | 568 | (5.6) | | 1 554 | (15.2) | 8 104 | (79.2) | | 2 153 | (21.1) | 7 505 | (73.4) |
| **Median age (IQR)** | 82 | (17) | 81 | (17) | | 74 | (19) | 83 | (16) | | 82 | (17) | 82 | (17) |
| **Average age (± SD)** | 78.8 | (14.8) | 77.1 | (16.1) | | 72.0 | (16.0) | 80.1 | (14.2) | | 78.7 | (15.0) | 79.2 | (13.9) |
| **N (%) females** | 4 909 | (50.8) | 266 | (46.8) | | 607 | (39.1) | 4 302 | (53.1) | | 1 091 | (50.7) | 3 818 | (50.9) |
| **N (%) external cause** | 598 | (6.2) | NA | NA | | 138 | (8.9) | 460 | (5.7) | | 106 | (4.9) | 492 | (6.6) |
| **N (%) death at home** | 1 554 | (16.1) | NA | NA | | 1 554 | (100.0) | 0 | (0.0) | | 280 | (13.0) | 1274 | (17.0) |
