## Supplementary table 3 for "Lockdown and non-COVID-19 deaths: Cause-specific mortality during the first wave of the 2020 pandemic in Norway. A population-based register study"

**Supplementary file 3:** Underlying data. Cause specific deaths March to May 2010 to 2020 with rates recorded in the Norwegian Cause of Death Registry by 11 December 2020. The causes have been grouped according to the EU Shortlist for Causes of Disease. Level 1 corresponds to the 17 main chapter

**Deaths (N)**

| Chapter | Level | Description | Deaths 2010 | Deaths 2011 | Deaths 2012 | Deaths 2013 | Deaths 2014 | Deaths 2015 | Deaths 2016 | Deaths 2017 | Deaths 2018 | Deaths 2019 | Deaths 2020 |
|---|---|---|---|---|---|---|---|---|---|---|---|---|---|
| | | **All causes** | 10179 | 10233 | 10574 | 10344 | 10110 | 10114 | 9947 | 9921 | 10401 | 9943 | 9658 |
| | | **Diseases** | 9555 | 9607 | 9980 | 9674 | 9423 | 9535 | 9301 | 9311 | 9701 | 9285 | 9060 |
| **1** | **1** | **Infectious and parasitic diseases** | **238** | **226** | **256** | **263** | **216** | **254** | **230** | **240** | **262** | **252** | **219** |
| 1.1 | 2 | Tuberculosis | 5 | 9 | 6 | 6 | 5 | 4 | 4 | 2 | 4 | 6 | 3 |
| 1.2 | 2 | AIDS (HIV-disease) | 1 | 3 | 4 | 2 | 5 | 3 | 1 | 2 | 3 | 2 | 4 |
| 1.3 | 2 | Viral hepatitis | 2 | 4 | 1 | 4 | 5 | 6 | 5 | 3 | 3 | 3 | 6 |
| 1.4 | 2 | Other infectious and parasitic diseases | 230 | 210 | 245 | 251 | 201 | 241 | 220 | 233 | 252 | 241 | 206 |
| **2** | **1** | **Neoplasms** | **2733** | **2821** | **2812** | **2786** | **2793** | **2757** | **2717** | **2765** | **2792** | **2706** | **2695** |
| **2.1** | **2** | **Malignant neoplasms** | **2660** | **2757** | **2749** | **2724** | **2736** | **2697** | **2643** | **2718** | **2722** | **2635** | **2608** |
| *2.1.1* | *3* | *Malignant neoplasm of lip, oral cavity, pharynx* | *25* | *36* | *42* | *34* | *28* | *37* | *49* | *37* | *39* | *38* | *44* |
| *2.1.2* | *3* | *Malignant neoplasm of oesophagus* | *56* | *46* | *47* | *53* | *54* | *59* | *54* | *41* | *50* | *60* | *74* |
| *2.1.3* | *3* | *Malignant neoplasm of stomach* | *81* | *95* | *80* | *80* | *75* | *92* | *70* | *73* | *69* | *69* | *69* |
| *2.1.4* | *3* | *Malignant neoplasm of colon, rectum and anus* | *392* | *393* | *427* | *408* | *402* | *410* | *398* | *410* | *394* | *358* | *343* |
| *2.1.5* | *3* | *Malignant neoplasm of liver and intrahepatic bile ducts* | *59* | *70* | *53* | *46* | *59* | *76* | *62* | *73* | *88* | *73* | *82* |
| *2.1.6* | *3* | *Malignant neoplasm of pancreas* | *155* | *154* | *179* | *167* | *150* | *171* | *186* | *182* | *189* | *191* | *187* |
| *2.1.7* | *3* | *Malignant neoplasm of larynx* | *11* | *10* | *12* | *13* | *10* | *7* | *4* | *8* | *11* | *5* | *7* |
| *2.1.8* | *3* | *Malignant neoplasm of trachea, bronchus, lung* | *548* | *584* | *573* | *588* | *556* | *544* | *536* | *538* | *555* | *556* | *543* |
| *2.1.9* | *3* | *Malignant melanoma of skin* | *81* | *79* | *78* | *77* | *95* | *82* | *71* | *83* | *75* | *66* | *73* |
| *2.1.10* | *3* | *Malignant neoplasm of breast* | *153* | *160* | *161* | *158* | *155* | *147* | *166* | *157* | *163* | *149* | *142* |
| *2.1.11* | *3* | *Malignant neoplasm of cervix uteri* | *20* | *9* | *19* | *15* | *16* | *20* | *25* | *22* | *27* | *19* | *20* |
| *2.1.12* | *3* | *Malignant neoplasm of other and unspecified parts of uterus* | *34* | *36* | *38* | *45* | *50* | *27* | *39* | *36* | *26* | *36* | *24* |
| *2.1.13* | *3* | *Malignant neoplasm of ovary* | *68* | *75* | *78* | *76* | *61* | *71* | *72* | *87* | *73* | *65* | *57* |
| *2.1.14* | *3* | *Malignant neoplasm of prostate* | *241* | *277* | *235* | *256* | *295* | *246* | *249* | *230* | *241* | *191* | *222* |
| *2.1.15* | *3* | *Malignant neoplasm of kidney* | *66* | *69* | *63* | *71* | *85* | *55* | *66* | *60* | *57* | *63* | *62* |
| *2.1.16* | *3* | *Malignant neoplasm of bladder* | *76* | *81* | *73* | *84* | *70* | *71* | *68* | *74* | *86* | *84* | *74* |
| *2.1.17* | *3* | *Malignant neoplasm of brain and central nervous system* | *91* | *74* | *69* | *72* | *81* | *86* | *75* | *81* | *81* | *85* | *80* |
| *2.1.18* | *3* | *Malignant neoplasm of thyroid* | *5* | *5* | *6* | *0* | *20* | *11* | *3* | *11* | *10* | *5* | *8* |
| *2.1.19* | *3* | *Hodgkin disease and lymphomas* | *69* | *101* | *80* | *93* | *77* | *89* | *62* | *81* | *67* | *73* | *89* |
| *2.1.20* | *3* | *Leukaemia* | *73* | *72* | *99* | *95* | *67* | *91* | *66* | *107* | *71* | *109* | *73* |
| *2.1.21* | *3* | *Other malignant neoplasm of lymphoid and haematopoietic tissue* | *72* | *67* | *73* | *63* | *73* | *66* | *65* | *62* | *71* | *72* | *57* |
| *2.1.22* | *3* | *Other malignant neoplasms* | *284* | *264* | *264* | *230* | *257* | *239* | *257* | *265* | *279* | *268* | *278* |
| 2.2 | 2 | Non-malignant neoplasms (benign and uncertain) | 73 | 64 | 63 | 62 | 57 | 60 | 74 | 47 | 70 | 71 | 87 |
| **3** | **1** | **Diseases of the blood and blood-forming organs and certain disorders involving the immune mechanism** | **45** | **40** | **34** | **30** | **38** | **32** | **32** | **29** | **37** | **33** | **36** |
| **4** | **1** | **Endocrine, nutritional and metabolic diseases** | **246** | **246** | **276** | **248** | **220** | **246** | **235** | **244** | **242** | **256** | **295** |
| 4.1 | 2 | Diabetes mellitus | 168 | 170 | 185 | 168 | 146 | 150 | 145 | 148 | 126 | 142 | 185 |
| 4.2 | 2 | Other endocrine, nutritional and metabolic diseases | 78 | 76 | 91 | 80 | 74 | 96 | 90 | 96 | 116 | 114 | 110 |
| **5** | **1** | **Mental and behavioural disorders** | **510** | **486** | **559** | **542** | **588** | **706** | **647** | **726** | **833** | **746** | **775** |
| 5.1 | 2 | Dementia | 417 | 411 | 490 | 487 | 527 | 619 | 576 | 659 | 750 | 695 | 697 |
| 5.2 | 2 | Alcohol abuse (including alcoholic psychosis) | 57 | 45 | 36 | 32 | 32 | 42 | 31 | 26 | 48 | 22 | 35 |
| 5.3 | 2 | Drug dependence, toxicomania | 6 | 6 | 6 | 4 | 3 | 8 | 2 | 2 | 8 | 2 | 7 |
| 5.4 | 2 | Other mental and behavioural disorders | 30 | 24 | 27 | 19 | 26 | 37 | 38 | 39 | 27 | 27 | 36 |
| **6** | **1** | **Diseases of the nervous system and the sense organs** | **406** | **373** | **433** | **438** | **479** | **497** | **482** | **530** | **552** | **584** | **625** |
| 6.1 | 2 | Parkinson's disease | 64 | 52 | 62 | 87 | 92 | 109 | 98 | 118 | 129 | 111 | 119 |
| 6.2 | 2 | Alzheimer's disease | 173 | 184 | 195 | 191 | 219 | 235 | 205 | 262 | 263 | 281 | 323 |
| 6.3 | 3 | Other diseases of the nervous system and the sense organs | 169 | 137 | 176 | 160 | 168 | 153 | 179 | 150 | 160 | 192 | 183 |
| **7** | **1** | **Diseases of the circulatory system** | **3350** | **3368** | **3346** | **3190** | **3013** | **2946** | **2860** | **2577** | **2624** | **2473** | **2182** |
| 7.1 | 2 | **Ischaemic heart diseases** | 1336 | 1291 | 1269 | 1206 | 1041 | 1041 | 1000 | 906 | 869 | 784 | 713 |

| Chapter | Level | Description | Deaths 2010 | Deaths 2011 | Deaths 2012 | Deaths 2013 | Deaths 2014 | Deaths 2015 | Deaths 2016 | Deaths 2017 | Deaths 2018 | Deaths 2019 | Deaths 2020 |
|---|---|---|---|---|---|---|---|---|---|---|---|---|---|
| *7.1.1* | *3* | *Acute myocardial infarction* | *839* | *843* | *794* | *757* | *656* | *633* | *595* | *515* | *523* | *447* | *380* |
| *7.1.2* | *2* | *Other ischaemic heart diseases* | *497* | *448* | *475* | *449* | *385* | *408* | *405* | *391* | *346* | *337* | *333* |
| 7.2 | 2 | Other heart diseases | 850 | 870 | 914 | 921 | 893 | 850 | 902 | 763 | 837 | 789 | 622 |
| 7.3 | 2 | Cerebrovascular diseases | 808 | 817 | 802 | 708 | 715 | 689 | 628 | 572 | 542 | 551 | 529 |
| 7.4 | 2 | Other diseases of the circulatory system | 356 | 390 | 361 | 355 | 364 | 366 | 330 | 336 | 376 | 349 | 318 |
| **8** | **1** | **Diseases of the respiratory system (excl. COVID-19)** | **977** | **1018** | **1188** | **1065** | **1010** | **1101** | **1114** | **1185** | **1299** | **1199** | **1005** |
| ***** | ***** | **Diseases of the respiratory system (incl. COVID-19)** | **977** | **1018** | **1188** | **1065** | **1010** | **1101** | **1114** | **1185** | **1299** | **1199** | **1221** |
| 8.1 | 2 | Influenza | 0 | 8 | 45 | 20 | 25 | 65 | 28 | 42 | 123 | 84 | 35 |
| 8.2 | 3 | Pneumonia | 391 | 382 | 445 | 382 | 333 | 367 | 405 | 456 | 398 | 376 | 302 |
| **8.3** | **2** | Chronic lower respiratory diseases | **496** | **534** | **581** | **555** | **535** | **547** | **572** | **582** | **648** | **621** | **541** |
| *8.3.1* | *3* | *Asthma* | *22* | *23* | *19* | *23* | *20* | *17* | *23* | *29* | *28* | *18* | *32* |
| *8.3.2* | *2* | *Other chronic lower respiratory diseases* | *474* | *511* | *562* | *532* | *515* | *530* | *549* | *553* | *620* | *603* | *509* |
| 8.4 | 2 | Other diseases of the respiratory system | 90 | 94 | 117 | 108 | 117 | 122 | 109 | 105 | 130 | 118 | 127 |
| **9** | **1** | **Diseases of the digestive system** | **302** | **341** | **324** | **313** | **313** | **301** | **304** | **337** | **316** | **336** | **302** |
| 9.1 | 2 | Ulcer of stomach, duodenum and jejunum | 48 | 43 | 50 | 44 | 36 | 40 | 43 | 34 | 38 | 31 | 21 |
| 9.2 | 2 | Cirrhosis, fibrosis and chronic hepatitis | 56 | 43 | 42 | 55 | 46 | 40 | 57 | 62 | 46 | 57 | 50 |
| 9.3 | 1 | Other diseases of the digestive system | 198 | 255 | 232 | 214 | 231 | 221 | 204 | 241 | 232 | 248 | 231 |
| **10** | **2** | **Diseases of the skin and subcutaneous tissue** | **15** | **22** | **18** | **20** | **24** | **32** | **31** | **38** | **27** | **28** | **27** |
| **11** | **1** | **Diseases of the musculoskeletal system/connective tissue** | **51** | **61** | **63** | **56** | **66** | **70** | **79** | **74** | **96** | **76** | **76** |
| 11.1 | 2 | Rheumatoid arthritis and osteoarthrosis | 11 | 16 | 11 | 13 | 16 | 14 | 23 | 21 | 33 | 19 | 15 |
| 11.2 | 2 | Other diseases of the musculoskeletal system/connective tissue | 40 | 45 | 52 | 43 | 50 | 56 | 56 | 53 | 63 | 57 | 61 |
| **12** | **1** | **Diseases of the genitourinary system** | **252** | **212** | **206** | **213** | **203** | **194** | **188** | **232** | **223** | **198** | **191** |
| 12.1 | 2 | Diseases of kidney and ureter | 165 | 130 | 127 | 129 | 112 | 122 | 113 | 150 | 135 | 125 | 131 |
| 12.2 | 1 | Other diseases of the genitourinary system | 87 | 82 | 79 | 84 | 91 | 72 | 75 | 82 | 88 | 73 | 60 |
| **13** | **1** | **Complications of pregnancy, childbirth and puerperium** | **0** | **1** | **0** | **1** | **0** | **0** | **0** | **0** | **1** | **0** | **0** |
| **14** | **1** | **Certain conditions originating in the perinatal period** | **28** | **21** | **27** | **28** | **22** | **24** | **14** | **16** | **15** | **8** | **22** |
| **15** | **2** | **Congenital malformations and chromosomal abnormalities** | **36** | **26** | **32** | **29** | **37** | **34** | **29** | **31** | **35** | **34** | **37** |
| **16** | **1** | **Symptoms, signs, ill-defined causes** | **366** | **345** | **406** | **452** | **401** | **341** | **339** | **287** | **347** | **356** | **357** |
| 16.1 | 2 | Sudden infant death syndrome | 2 | 2 | 5 | 1 | 2 | 6 | 3 | 1 | 0 | 2 | 0 |
| 16.2 | 2 | Unknown and unspecified causes | 231 | 237 | 242 | 319 | 270 | 216 | 197 | 170 | 225 | 260 | 254 |
| 16.3 | 2 | Other symptoms, signs, ill-defined causes | 133 | 106 | 159 | 132 | 129 | 119 | 139 | 116 | 122 | 94 | 103 |
| **17** | **1** | **External causes of morbidity and mortality** | **624** | **626** | **594** | **670** | **687** | **579** | **646** | **610** | **700** | **658** | **598** |
| 17.1 | 2 | Accidents | 459 | 458 | 447 | 498 | 519 | 429 | 481 | 446 | 488 | 485 | 452 |
| *17.1.1* | *3* | *Transport accidents* | *54* | *50* | *42* | *47* | *44* | *40* | *46* | *34* | *30* | *38* | *35* |
| *17.1.2* | *3* | *Accidental falls* | *89* | *104* | *106* | *124* | *138* | *202* | *216* | *151* | *177* | *165* | *161* |
| *17.1.3* | *3* | *Drowning and accidental submersion* | *17* | *15* | *8* | *8* | *19* | *10* | *12* | *15* | *19* | *7* | *13* |
| *17.1.4* | *3* | *Accidental poisoning* | *79* | *71* | *78* | *87* | *80* | *78* | *102* | *81* | *94* | *90* | *79* |
| *17.1.5* | *3* | *Other accidents* | *220* | *218* | *213* | *232* | *238* | *99* | *105* | *165* | *168* | *185* | *164* |
| 17.2 | 2 | Suicide and intentional self-harm | 161 | 156 | 142 | 159 | 158 | 144 | 156 | 162 | 206 | 166 | 139 |
| 17.3 | 2 | Homicide, assault | 4 | 9 | 3 | 13 | 5 | 6 | 8 | 1 | 4 | 4 | 6 |
| 17.4 | 2 | Events of undetermined intent | 0 | 1 | 1 | 0 | 5 | 0 | 1 | 1 | 2 | 1 | 0 |
| 17.5 | 2 | Other external causes of injury and poisoning | 0 | 2 | 1 | 0 | 0 | 0 | 0 | 0 | 0 | 2 | 1 |
|  | **1** | **COVID-19** | **0** | **0** | **0** | **0** | **0** | **0** | **0** | **0** | **0** | **0** | **216** |
|  |  | Missing | 164 | 151 | 131 | 165 | 147 | 156 | 164 | 176 | 169 | 295 | 568 |
|  | **0** | **Total** | **10343** | **10384** | **10705** | **10509** | **10257** | **10270** | **10111** | **10097** | **10570** | **10238** | **10226** |

**Death rates** 2010-2020. Age-standardised (EU 2013 population) and adjusted for missing causes of death.

| Chapter | Level | Description | Rate 2010 | Rate 2011 | Rate 2012 | Rate 2013 | Rate 2014 | Rate 2015 | Rate 2016 | Rate 2017 | Rate 2018 | Rate 2019 | Rate 2020 |
|---|---|---|---|---|---|---|---|---|---|---|---|---|---|
| | | **All causes** | 251.56 | 248.37 | 253.07 | 244.06 | 235.52 | 231.72 | 223.72 | 219.36 | 226.59 | 212.8 | 202.56 |
| | | **Diseases** | 237.35 | 234.28 | 239.98 | 229.35 | 220.6 | 219.38 | 210.12 | 206.69 | 212.11 | 199.41 | 190.49 |
| **1** | **1** | **Infectious and parasitic diseases** | **5.77** | **5.44** | **6.02** | **6.18** | **4.95** | **5.78** | **5.22** | **5.32** | **5.76** | **5.5** | **4.64** |
| 1.1 | 2 | Tuberculosis | 0.12 | 0.21 | 0.12 | 0.15 | 0.12 | 0.09 | 0.09 | 0.05 | 0.09 | 0.13 | 0.06 |
| 1.2 | 2 | AIDS (HIV-disease) | 0.02 | 0.07 | 0.09 | 0.04 | 0.09 | 0.07 | 0.02 | 0.04 | 0.07 | 0.04 | 0.08 |
| 1.3 | 2 | Viral hepatitis | 0.04 | 0.09 | 0.02 | 0.08 | 0.1 | 0.12 | 0.1 | 0.06 | 0.06 | 0.06 | 0.12 |
| 1.4 | 2 | Other infectious and parasitic diseases | 5.59 | 5.07 | 5.79 | 5.91 | 4.64 | 5.5 | 5.01 | 5.17 | 5.54 | 5.27 | 4.38 |
| **2** | **1** | **Neoplasms** | **68.28** | **69.55** | **68.22** | **66.1** | **65.85** | **63.52** | **61.33** | **61.04** | **60.78** | **57.57** | **56.13** |
| **2.1** | **2** | **Malignant neoplasms** | **66.48** | **67.98** | **66.62** | **64.65** | **64.49** | **62.14** | **59.65** | **59.99** | **59.26** | **56.03** | **54.29** |
| *2.1.1* | *3* | *Malignant neoplasm of lip, oral cavity, pharynx* | *0.64* | *0.88* | *0.99* | *0.81* | *0.65* | *0.81* | *1.09* | *0.82* | *0.82* | *0.79* | *0.92* |
| *2.1.2* | *3* | *Malignant neoplasm of oesophagus* | *1.37* | *1.17* | *1.1* | *1.29* | *1.28* | *1.35* | *1.24* | *0.88* | *1.1* | *1.25* | *1.51* |
| *2.1.3* | *3* | *Malignant neoplasm of stomach* | *1.98* | *2.33* | *1.97* | *1.87* | *1.79* | *2.09* | *1.61* | *1.61* | *1.49* | *1.47* | *1.44* |
| *2.1.4* | *3* | *Malignant neoplasm of colon, rectum and anus* | *9.87* | *9.67* | *10.38* | *9.69* | *9.46* | *9.44* | *8.95* | *9.11* | *8.63* | *7.57* | *7.15* |
| *2.1.5* | *3* | *Malignant neoplasm of liver and intrahepatic bile ducts* | *1.46* | *1.65* | *1.27* | *1.08* | *1.38* | *1.74* | *1.35* | *1.62* | *1.88* | *1.56* | *1.69* |
| *2.1.6* | *3* | *Malignant neoplasm of pancreas* | *3.9* | *3.79* | *4.41* | *4.06* | *3.56* | *3.97* | *4.27* | *4.02* | *4.16* | *4.09* | *3.89* |
| *2.1.7* | *3* | *Malignant neoplasm of larynx* | *0.28* | *0.25* | *0.3* | *0.33* | *0.24* | *0.16* | *0.09* | *0.17* | *0.24* | *0.11* | *0.14* |
| *2.1.8* | *3* | *Malignant neoplasm of trachea, bronchus, lung* | *13.86* | *14.68* | *14.07* | *14.06* | *13.15* | *12.71* | *12.18* | *11.94* | *12.11* | *11.73* | *11.27* |
| *2.1.9* | *3* | *Malignant melanoma of skin* | *1.98* | *1.92* | *1.88* | *1.81* | *2.22* | *1.8* | *1.56* | *1.82* | *1.61* | *1.4* | *1.52* |
| *2.1.10* | *3* | *Malignant neoplasm of breast* | *3.82* | *3.82* | *3.86* | *3.62* | *3.5* | *3.31* | *3.63* | *3.38* | *3.47* | *3.12* | *2.93* |
| *2.1.11* | *3* | *Malignant neoplasm of cervix uteri* | *0.45* | *0.23* | *0.41* | *0.34* | *0.36* | *0.44* | *0.55* | *0.44* | *0.56* | *0.38* | *0.4* |
| *2.1.12* | *3* | *Malignant neoplasm of other and unspecified parts of uterus* | *0.85* | *0.89* | *0.92* | *1.08* | *1.17* | *0.63* | *0.88* | *0.82* | *0.57* | *0.79* | *0.5* |
| *2.1.13* | *3* | *Malignant neoplasm of ovary* | *1.7* | *1.8* | *1.79* | *1.77* | *1.43* | *1.61* | *1.61* | *1.91* | *1.56* | *1.38* | *1.16* |
| *2.1.14* | *3* | *Malignant neoplasm of prostate* | *6.06* | *6.91* | *5.72* | *6.17* | *7.08* | *5.74* | *5.77* | *5.16* | *5.3* | *4.11* | *4.75* |
| *2.1.15* | *3* | *Malignant neoplasm of kidney* | *1.67* | *1.67* | *1.54* | *1.7* | *2.02* | *1.29* | *1.48* | *1.31* | *1.23* | *1.33* | *1.28* |
| *2.1.16* | *3* | *Malignant neoplasm of bladder* | *1.89* | *2.02* | *1.81* | *2.01* | *1.61* | *1.66* | *1.57* | *1.64* | *1.91* | *1.84* | *1.55* |
| *2.1.17* | *3* | *Malignant neoplasm of brain and central nervous system* | *2.16* | *1.74* | *1.55* | *1.62* | *1.81* | *1.88* | *1.63* | *1.71* | *1.71* | *1.75* | *1.59* |
| *2.1.18* | *3* | *Malignant neoplasm of thyroid* | *0.13* | *0.13* | *0.15* | *0* | *0.45* | *0.25* | *0.07* | *0.24* | *0.22* | *0.1* | *0.17* |
| *2.1.19* | *3* | *Hodgkin disease and lymphomas* | *1.69* | *2.56* | *1.94* | *2.15* | *1.86* | *2.1* | *1.4* | *1.85* | *1.49* | *1.6* | *1.87* |
| *2.1.20* | *3* | *Leukaemia* | *1.85* | *1.76* | *2.44* | *2.21* | *1.55* | *2.06* | *1.48* | *2.35* | *1.56* | *2.37* | *1.54* |
| *2.1.21* | *3* | *Other malignant neoplasm of lymphoid and haematopoietic tissue* | *1.83* | *1.67* | *1.81* | *1.46* | *1.78* | *1.52* | *1.47* | *1.37* | *1.57* | *1.55* | *1.19* |
| *2.1.22* | *3* | *Other malignant neoplasms* | *7.04* | *6.44* | *6.31* | *5.52* | *6.14* | *5.58* | *5.77* | *5.82* | *6.07* | *5.74* | *5.83* |
| 2.2 | 2 | Non-malignant neoplasms (benign and uncertain) | 1.8 | 1.57 | 1.6 | 1.45 | 1.36 | 1.38 | 1.68 | 1.05 | 1.52 | 1.54 | 1.84 |
| **3** | **1** | **Diseases of the blood and blood-forming organs and certain disorders involving the immune mechanism** | **1.13** | **1.02** | **0.81** | **0.71** | **0.86** | **0.75** | **0.71** | **0.64** | **0.81** | **0.69** | **0.76** |
| **4** | **1** | **Endocrine, nutritional and metabolic diseases** | **6.04** | **5.9** | **6.58** | **5.8** | **5.11** | **5.65** | **5.34** | **5.41** | **5.22** | **5.49** | **6.18** |
| 4.1 | 2 | Diabetes mellitus | 4.13 | 4.08 | 4.4 | 3.93 | 3.4 | 3.48 | 3.3 | 3.29 | 2.73 | 3.06 | 3.92 |
| 4.2 | 2 | Other endocrine, nutritional and metabolic diseases | 1.91 | 1.82 | 2.18 | 1.87 | 1.71 | 2.17 | 2.04 | 2.12 | 2.49 | 2.43 | 2.26 |
| **5** | **1** | **Mental and behavioural disorders** | **12.39** | **11.73** | **13.27** | **12.9** | **13.54** | **16.18** | **14.54** | **16.14** | **18.26** | **16.22** | **16.55** |
| 5.1 | 2 | Dementia | 10.27 | 10 | 11.67 | 11.66 | 12.2 | 14.27 | 12.96 | 14.7 | 16.5 | 15.14 | 14.94 |
| 5.2 | 2 | Alcohol abuse (including alcoholic psychosis) | 1.28 | 1.06 | 0.82 | 0.7 | 0.7 | 0.92 | 0.67 | 0.54 | 1.02 | 0.46 | 0.71 |
| 5.3 | 2 | Drug dependence, toxicomania | 0.12 | 0.13 | 0.13 | 0.08 | 0.06 | 0.16 | 0.05 | 0.04 | 0.15 | 0.04 | 0.13 |
| 5.4 | 2 | Other mental and behavioural disorders | 0.72 | 0.54 | 0.65 | 0.46 | 0.58 | 0.83 | 0.86 | 0.86 | 0.59 | 0.58 | 0.77 |
| **6** | **1** | **Diseases of the nervous system and the sense organs** | **10.1** | **9.14** | **10.37** | **10.44** | **11.4** | **11.53** | **10.96** | **11.9** | **12.1** | **12.58** | **13.22** |
| 6.1 | 2 | Parkinson's disease | 1.64 | 1.27 | 1.5 | 2.11 | 2.24 | 2.6 | 2.3 | 2.75 | 2.91 | 2.46 | 2.54 |
| 6.2 | 2 | Alzheimer's disease | 4.26 | 4.52 | 4.73 | 4.59 | 5.21 | 5.5 | 4.72 | 5.92 | 5.82 | 6.15 | 6.94 |
| 6.3 | 3 | Other diseases of the nervous system and the sense organs | 4.2 | 3.35 | 4.14 | 3.74 | 3.95 | 3.43 | 3.94 | 3.23 | 3.37 | 3.97 | 3.74 |
| **7** | **1** | **Diseases of the circulatory system** | **83.49** | **81.77** | **80.39** | **75.66** | **70.43** | **67.77** | **64.63** | **57.32** | **57.39** | **53.33** | **46.03** |
| 7.1 | 2 | **Ischaemic heart diseases** | 33.47 | 31.44 | 30.33 | 28.41 | 24.41 | 23.99 | 22.54 | 20.12 | 18.99 | 16.76 | 14.99 |
| *7.1.1* | *3* | *Acute myocardial infarction* | *21.03* | *20.51* | *18.91* | *17.82* | *15.45* | *14.57* | *13.41* | *11.44* | *11.48* | *9.59* | *8.02* |
| *7.1.2* | *2* | *Other ischaemic heart diseases* | *12.44* | *10.93* | *11.42* | *10.59* | *8.96* | *9.42* | *9.13* | *8.68* | *7.51* | *7.17* | *6.97* |
| 7.2 | 2 | Other heart diseases | 21.11 | 21.03 | 21.84 | 21.72 | 20.72 | 19.43 | 20.36 | 16.89 | 18.23 | 17.07 | 13.2 |

| Chapter | Level | Description | Rate 2010 | Rate 2011 | Rate 2012 | Rate 2013 | Rate 2014 | Rate 2015 | Rate 2016 | Rate 2017 | Rate 2018 | Rate 2019 | Rate 2020 |
|---|---|---|---|---|---|---|---|---|---|---|---|---|---|
| 7.3 | 2 | Cerebrovascular diseases | 19.94 | 19.78 | 19.43 | 17.02 | 16.67 | 15.84 | 14.31 | 12.82 | 11.94 | 11.98 | 11.17 |
| 7.4 | 2 | Other diseases of the circulatory system | 8.97 | 9.52 | 8.79 | 8.51 | 8.63 | 8.51 | 7.42 | 7.49 | 8.23 | 7.52 | 6.67 |
| 8 | 1 | Diseases of the respiratory system (excl. COVID-19) | 24.52 | 25.16 | 28.97 | 25.65 | 24.02 | 25.7 | 25.45 | 26.61 | 28.75 | 25.89 | 21.29 |
| * | * | Diseases of the respiratory system (incl. COVID-19) | 24,52 | 25,16 | 28,97 | 25,65 | 24,02 | 25,7 | 25,45 | 26,61 | 28,75 | 25,89 | 25,93 |
| 8.1 | 2 | Influenza | 0 | 0.21 | 1.13 | 0.49 | 0.57 | 1.49 | 0.63 | 0.95 | 2.72 | 1.81 | 0.72 |
| 8.2 | 3 | Pneumonia | 9.71 | 9.41 | 10.63 | 9.02 | 7.77 | 8.48 | 9.09 | 10.1 | 8.72 | 8.11 | 6.4 |
| 8.3 | 2 | Chronic lower respiratory diseases | 12.6 | 13.24 | 14.34 | 13.57 | 12.87 | 12.88 | 13.25 | 13.2 | 14.45 | 13.45 | 11.47 |
| 8.3.1 | 3 | Asthma | 0.53 | 0.56 | 0.45 | 0.54 | 0.45 | 0.39 | 0.52 | 0.63 | 0.61 | 0.39 | 0.68 |
| 8.3.2 | 2 | Other chronic lower respiratory diseases | 12.07 | 12.68 | 13.89 | 13.03 | 12.42 | 12.49 | 12.73 | 12.57 | 13.84 | 13.06 | 10.79 |
| 8.4 | 2 | Other diseases of the respiratory system | 2.21 | 2.3 | 2.87 | 2.57 | 2.81 | 2.85 | 2.48 | 2.36 | 2.86 | 2.52 | 2.7 |
| 9 | 1 | Diseases of the digestive system | 7.38 | 8.29 | 7.73 | 7.37 | 7.32 | 6.9 | 6.84 | 7.41 | 6.86 | 7.22 | 6.34 |
| 9.1 | 2 | Ulcer of stomach, duodenum and jejunum | 1.15 | 1.04 | 1.19 | 1.05 | 0.84 | 0.92 | 0.96 | 0.76 | 0.82 | 0.65 | 0.43 |
| 9.2 | 2 | Cirrhosis, fibrosis and chronic hepatitis | 1.36 | 1.03 | 1 | 1.23 | 1.03 | 0.88 | 1.25 | 1.32 | 0.97 | 1.17 | 1.02 |
| 9.3 | 1 | Other diseases of the digestive system | 4.87 | 6.22 | 5.54 | 5.09 | 5.45 | 5.1 | 4.63 | 5.33 | 5.07 | 5.4 | 4.89 |
| 10 | 2 | Diseases of the skin and subcutaneous tissue | 0.36 | 0.54 | 0.45 | 0.46 | 0.56 | 0.75 | 0.7 | 0.84 | 0.6 | 0.62 | 0.58 |
| 11 | 1 | Diseases of the musculoskeletal system/connective tissue | 1.3 | 1.49 | 1.47 | 1.31 | 1.53 | 1.63 | 1.79 | 1.67 | 2.14 | 1.62 | 1.6 |
| 11.1 | 2 | Rheumatoid arthritis and osteoarthrosis | 0.29 | 0.38 | 0.25 | 0.31 | 0.37 | 0.33 | 0.53 | 0.49 | 0.75 | 0.4 | 0.32 |
| 11.2 | 2 | Other diseases of the musculoskeletal system/connective tissue | 1.01 | 1.11 | 1.22 | 1 | 1.16 | 1.3 | 1.26 | 1.18 | 1.39 | 1.22 | 1.28 |
| 12 | 1 | Diseases of the genitourinary system | 6.17 | 5.08 | 4.89 | 5.07 | 4.71 | 4.44 | 4.25 | 5.22 | 4.97 | 4.29 | 4.04 |
| 12.1 | 2 | Diseases of kidney and ureter | 4.03 | 3.13 | 3.04 | 3.08 | 2.61 | 2.76 | 2.54 | 3.38 | 3 | 2.74 | 2.77 |
| 12.2 | 1 | Other diseases of the genitourinary system | 2.14 | 1.95 | 1.85 | 1.99 | 2.1 | 1.68 | 1.71 | 1.84 | 1.97 | 1.55 | 1.27 |
| 13 | 1 | Complications of pregnancy, childbirth and puerperium | 0 | 0.02 | 0 | 0.02 | 0 | 0 | 0 | 0 | 0.02 | 0 | 0 |
| 14 | 1 | Certain conditions originating in the perinatal period | 0.46 | 0.34 | 0.45 | 0.46 | 0.37 | 0.4 | 0.24 | 0.27 | 0.26 | 0.14 | 0.4 |
| 15 | 2 | Congenital malformations and chromosomal abnormalities | 0.7 | 0.51 | 0.61 | 0.56 | 0.73 | 0.65 | 0.56 | 0.58 | 0.65 | 0.65 | 0.71 |
| 16 | 1 | Symptoms, signs, ill-defined causes | 9.26 | 8.3 | 9.75 | 10.66 | 9.22 | 7.73 | 7.56 | 6.32 | 7.54 | 7.6 | 7.38 |
| 16.1 | 2 | Sudden infant death syndrome | 0.03 | 0.03 | 0.08 | 0.02 | 0.03 | 0.1 | 0.05 | 0.02 | 0 | 0.04 | 0 |
| 16.2 | 2 | Unknown and unspecified causes | 5.77 | 5.62 | 5.77 | 7.48 | 6.18 | 4.91 | 4.41 | 3.77 | 4.93 | 5.55 | 5.24 |
| 16.3 | 2 | Other symptoms, signs, ill-defined causes | 3.46 | 2.65 | 3.9 | 3.16 | 3.01 | 2.72 | 3.1 | 2.53 | 2.61 | 2.01 | 2.14 |
| 17 | 1 | External causes of morbidity and mortality | 14.21 | 14.09 | 13.09 | 14.71 | 14.92 | 12.34 | 13.6 | 12.67 | 14.48 | 13.39 | 12.07 |
| 17.1 | 2 | Accidents | 10.65 | 10.5 | 10.09 | 11.17 | 11.52 | 9.37 | 10.34 | 9.52 | 10.41 | 10.06 | 9.32 |
| 17.1.1 | 3 | Transport accidents | 1.16 | 1.03 | 0.86 | 0.96 | 0.92 | 0.84 | 0.93 | 0.69 | 0.58 | 0.75 | 0.67 |
| 17.1.2 | 3 | Accidental falls | 2.19 | 2.55 | 2.49 | 2.91 | 3.17 | 4.61 | 4.83 | 3.36 | 3.92 | 3.52 | 3.46 |
| 17.1.3 | 3 | Drowning and accidental submersion | 0.37 | 0.31 | 0.19 | 0.16 | 0.39 | 0.2 | 0.25 | 0.29 | 0.4 | 0.13 | 0.25 |
| 17.1.4 | 3 | Accidental poisoning | 1.65 | 1.45 | 1.57 | 1.72 | 1.58 | 1.52 | 2.02 | 1.57 | 1.81 | 1.71 | 1.5 |
| 17.1.5 | 3 | Other accidents | 5.28 | 5.16 | 4.98 | 5.42 | 5.46 | 2.2 | 2.31 | 3.61 | 3.7 | 3.95 | 3.44 |
| 17.2 | 2 | Suicide and intentional self-harm | 3.48 | 3.34 | 2.9 | 3.28 | 3.2 | 2.85 | 3.07 | 3.11 | 3.96 | 3.18 | 2.62 |
| 17.3 | 2 | Homicide, assault | 0.08 | 0.19 | 0.06 | 0.26 | 0.09 | 0.12 | 0.17 | 0.02 | 0.07 | 0.08 | 0.11 |
| 17.4 | 2 | Events of undetermined intent | 0 | 0.02 | 0.02 | 0 | 0.11 | 0 | 0.02 | 0.02 | 0.04 | 0.02 | 0 |
| 17.5 | 2 | Other external causes of injury and poisoning | 0 | 0.04 | 0.02 | 0 | 0 | 0 | 0 | 0 | 0 | 0.05 | 0.02 |
| | 1 | COVID-19 | 0 | 0 | 0 | 0 | 0 | 0 | 0 | 0 | 0 | 0 | 4.64 |
| | | Missing | 3.98 | 3.54 | 3.04 | 3.87 | 3.42 | 3.47 | 3.53 | 3.78 | 3.61 | 6.28 | 11.83 |
| | 0 | Total | 255.53 | 251.86 | 256.13 | 247.91 | 238.98 | 235.24 | 227.24 | 223.11 | 230.21 | 219.11 | 214.38 |
